# Implementing the Maternal Postnatal Attachment Scale (MPAS) in universal services: Qualitative interviews with health visitors

**DOI:** 10.1101/2021.11.30.21267065

**Authors:** Philippa K Bird, Zoe Hindson, Abigail Dunn, Anna Cronin de Chavez, Josie Dickerson, Joanna Howes, Tracey Bywater

**Author notes:** Note: Philippa Bird’s current affiliation is Leeds Teaching Hospitals Trust, Leeds, UK Anna Cronin de Chavez’s current affiliation is London School of Hygiene and Tropical Medicine Joanna Howes’ current affiliation is Bradford Metropolitan District Council, Bradford, UK Zoe Hindson’s current affiliation is Family Action, UK. Corresponding author: Josie Dickerson, Bradford Institute for Health Research, Bradford Royal Infirmary, Duckworth Ln, Bradford BD9 6RJ. **Authorship/co-authorship statement:** PKB, AD, JD, ZH and TB designed the study, PKB, AD and AC conducted interviews, PKB, AC and ZH analysed the data collected. All authors interpreted findings, have contributed to the manuscript substantially and have agreed to the final submitted version.

## Abstract

A secure parent-infant relationship lays the foundations for children’s development, however there are currently no measurement tools recommended for clinical practice. We evaluate the clinical utility of a structured assessment of the parent-infant relationship (the Maternal Postnatal Attachment Scale, MPAS) in a deprived, multi-ethnic urban community in England. This paper answers the question: what are health visitors’ views on the parent-infant relationship, and experiences of piloting the MPAS? It explores the barriers and facilitators to implementation, and complements the paper on psychometric properties and representativeness reported in Dunn et al (submitted).

Semi-structured interviews were conducted with 11 health visitors and data were analysed using thematic analysis. Health visitors stressed the importance of the parent-infant relationship and reported benefits of the MPAS, including opening conversation, and identifying and reporting concerns. Challenges included timing, workload, the appropriateness and understanding of the questions and the length of the tool. Suggestions for improvements to the tool were identified.

Our findings help to explain results in Dunn et al, and challenges identified would hinder routine assessment of the parent-infant relationship. Further work with health professionals and parents has been undertaken to co-produce an acceptable, feasible and reliable tool for clinical practice.

**Key findings and points for practitioners:**

- Health visitors saw identification and support of the parent-infant relationship as an important part of their role, however there are currently no recommended tools for this.
- Health visitors report some benefits to using the MPAS, but also several challenges to using this tool in practice, including the length of time required, the complexity of the language, potential to trigger distress and perceived intrusiveness of some questions.
- Further work in collaboration with health professionals and parents is needed to develop an acceptable, feasible and reliable tool to assess the parent-infant relationship.

**Statement of relevance to the field of infant and early childhood mental health:** A secure parent-infant relationship lays the foundations for children’s development, and identification of concerns and provision of support is a priority in the UK and internationally. However, no tools are currently recommended for assessing the relationship in clinical practice. Our findings on experiences, benefits and challenges of piloting a tool to assess the parent-infant relationship provide important directions for development of a short, clinically relevant and valid tool in clinical practice.

**Statement explaining how the research reflects an appreciation for diversity and an anti-racist approach:** This pilot was conducted with a diverse, multi-ethnic community (half of new mothers are from Asian/Asian British Pakistani backgrounds, a quarter White British, and a quarter from other ethnic backgrounds). The health visiting service engages with the whole population in a culturally sensitive way, including ensuring staff speak key community languages and using interpreters. None of the authors spoke community languages, but we purposively selected the health visitors to include experience of using the MPAS in community languages. Our findings reflect experiences implementing the tool with women from different ethnic backgrounds, and we report detailed findings on language. We hope our findings can inform appropriate and equitable implementation of tools in diverse communities.

## Introduction

A secure parent-infant relationship lays the foundations for children’s socio-emotional development (Cassidy et al., 2013; Fearon et al., 2010; Fernald et al., 2013; Stams et al., 2002). This is increasingly recognised in policy and service provision guidelines in the UK and internationally (NICE, 2012, 2015, 2020). Early identification of, and intervention to support, mothers who are struggling to develop an appropriate relationship with their infant has been identified as a priority for health visiting in the UK (Public Health England, 2021). However, no tools are currently recommended for assessing the relationship between parents and infants under 12 months, a developmentally critical time period for attachment (NICE, 2015).

There are a number of different approaches to measuring the parent-infant relationship, including questionnaires to understand the maternal experience and observation of maternal and infant behaviour (Gridley et al., 2019; Lotzin et al., 2015). However, there is a lack of robust, measures validated in the UK, and recent reviews have concluded that none of the available measures are recommended for clinical use due to inadequate evidence on the psychometric properties and clinical utility (Mathews et al., 2019; Wittkowski et al., 2020). A detailed overview of these measurement approaches is provided in Dunn et al. (submitted).

In the UK, health visiting is a universal nursing service providing home visits to support mental health, parenting and infant development. Equivalent roles in other countries may be a public health nurse, or child and family health nurse. There is substantial variation in how health visitors assess the parent-infant relationship, with unstructured observation and professional judgement commonly used (Appleton et al., 2013; Wilson et al., 2010). This also means that no routine data are available to understand the prevalence of attachment concerns in the population and drive action. There is a need for a short, clinically relevant and valid tool for use in universal services (Nunes et al., 2014).

A pilot of implementation of the Maternal Postnatal Attachment Scale (MPAS), a structured quantitative assessment of the parent-infant relationship, took place in universal health visiting services in a deprived and ethnically diverse urban area. The pilot was part of Better Start Bradford, a 10-year programme funded by the National Lottery Community Fund to promote the health and development of children aged 0-3 in an area of Bradford, a city in the north of England. The majority of the pilot area is within the 10% most deprived areas in England and the population is ethnically diverse, with 49% of new mothers from Asian/Asian British Pakistani backgrounds, 25% White British, 5% Asian/Asian British Bangladeshi, 4% White other, and 18% from other ethnic backgrounds (Dickerson et al., 2016).

The MPAS (Condon & Corkindale, 1998) was selected by a team of clinical specialists and the evaluation team as the most appropriate tool based on a scoping/rapid review of the current literature and expected clinical utility. MPAS is a self-complete measure of maternal affect towards the infant, designed to be completed by mothers of infants between birth and 12 months. Further information on the tool is provided in Dunn et al. (In preparation).

The evaluation of the pilot assessed the clinical utility of the MPAS in routine health visiting care in a deprived, multi-ethnic urban community in Bradford, UK. This is the second of two linked papers describing the quantitative and qualitative findings. The first (Dunn et al., submitted) presented an introduction to MPAS and quantitative findings on the measure’s psychometric properties and the coverage of the tool amongst the eligible population. Findings showed that the tool has limited validity for assessing mothers’ affective bond to their infant, suggesting that it is not robust enough to recommend ongoing use in the Bradford context.

This second paper reports the qualitative component of the evaluation, and further discussion of the findings from both papers. The aim of the qualitative study was to understand health visitors’ views on the parent-infant relationship and their experience of using MPAS, to develop recommendations for future use. This included understanding:

- How health visitors conceptualised the parent-infant relationship and their role in supporting it.
- Health visitor experiences of using the MPAS with mothers.
- Future service development needs to improve measurement of the parent-infant relationship, including training and use of the MPAS.

## Methods

### Background

Health visitors were asked to administer the MPAS to all mothers at the 3-4-month home visit as a pilot in an area of Bradford (May 2017-May 2018). Health visitors received training on the parent-infant relationship, administration of the MPAS and scoring responses at the start of the pilot. Mothers were asked to self-complete the MPAS, although health visitors supported completion where required. If they had limited understanding of English, a bilingual health visitor or interpreter administered the tool with the health visitor present.

### Participants

Semi-structured interviews were conducted with 11 female health visitors out of the 37 working in the pilot area. Health visitors were provided with study information, including an explanation that interviews were voluntary, via their team leaders. Health visitors were then approached to take part in an interview. The group were selected purposively to include those working in different parts of the pilot area, with varied levels of use of MPAS, and bilingual professionals (Urdu language). Immediately prior to the interview, the researcher provided further detailed information on the study and informed written consent was obtained.

### Interviews

The interview guide was piloted and amended prior to use (supplementary information). Interviews were conducted in English and lasted approximately 40 minutes (10 face-to-face, 1 telephone). Interviews were transcribed verbatim and thematic analysis was conducted to identify and report patterns within the data (Braun & Clarke, 2006).

### Analysis

The coding frame was developed (PB, ZH and AC) using emerging themes from the interviews, and a-priori themes from research questions and literature. Transcripts were coded systematically (PB and ZH) and findings analysed by theme. Transcripts and analysis were managed using NVivo 11 software.

### Ethics

The study followed a protocol. Health visitors received information in advance of being approached for an interview, followed by an informed consent process at the start of interviews. They were informed that participation was voluntary and that they could stop at any point. Confidentiality was protected through removal of names from the data prior to analysis. Data are stored securely at the Bradford Teaching Hospital. The Health Research Authority confirmed that this study is considered to be service evaluation, not research, and as such did not require review by an NHS Research Ethics Committee (HRA decision 60/88/81).

## Findings

*Five themes were identified, with corresponding sub-themes: 1) Health visitors’ role and the parent-infant relationship; 2) Assessment of the parent-infant relationship; 3) Benefits of using MPAS in clinical practice (with sub-themes of promoting dialogue; identifying and recording concerns); 4) Challenges of using MPAS in clinical practice (with subthemes of timing and workload, appropriateness and understanding of questions, length and repetition, scoring and referrals, and English competency); and 5) Suggestions to improve assessment and support of the parent-infant relationship*.

### 1) Health visitors’ role and the parent-infant relationship

During the interviews, health visitors stressed the importance of a healthy parent-infant relationship. Descriptions of what this meant included interaction with the baby, being attentive (including being ‘in tune’ and responsive parenting), and developing bonds and affection with the baby. Respondents commonly referred to a strong parent-infant relationship laying the foundations for child health and wellbeing:

> Well, I think it’s absolutely crucial you know, right from pregnancy obviously we promote it, but it’s crucial to get that parent relationship right really to give that infant, that baby, the best start in life (HV10)

Health visitors noted their role in facilitating a positive parent-infant relationship, and described approaches used including the provision of additional support (e.g. “listening visits”), information or signposting to other services. Some of the respondents felt that health visitors have a unique opportunity to assess and support the parent-infant relationship, as home visits help understand the wider family context:

> I think we have lots of influence because when we go into the family home, we’re not just assessing what that parent-infant relationship is in isolation, we’re looking at everything. The parent-infant relationship might be strained but that might be because of drug use or finances or domestic abuse or lots of things. So, by assessing where areas of concern are and putting early interventions into place… we’re indirectly influencing that relationship (HV8)

### 2) Assessment of the parent-infant relationship

Assessment of the parent-infant relationship was generally considered an important part of practice. Health visitors stressed the importance of observation and listening, describing how careful observation of body language, eye contact, and feeding provides insights:

> I think observation, it’s got to be observation…. Yes, parents sometimes tell us how they are feeling, but sometimes it’s the non-verbal communication that is slightly better, it’s the way they behave, the way they handle, the way they look at the baby (HV1)

Several respondents advocated for open or interactive discussions rather than a questionnaire, due to concerns that mothers would answer structured questions as they believe they should, rather than honestly. Training, experience and the development of a strong relationship with mothers were considered key enablers for observation and discussion.

All respondents reported using MPAS to some extent, ranging from some, to all, 3-4-month contacts. Reasons given for not using it every time included the mother being unwilling, insufficient time, or inappropriateness given the circumstances. Several health visitors suggested that MPAS should be used only after building a trusting relationship:

> I wouldn’t want to use this with a family in… a busy clinic when I’ve never met that family before. I think it’s something that you use alongside with your relationship building skills and it’s to be used at a time when the family trust you and [are] able to discuss their feelings (HV4)

### 3) Benefits of using MPAS in clinical practice

#### Promoting dialogue

MPAS was viewed by many as a useful conversation-starter around the parent-infant relationship. Some indicated the conversation prompted by using MPAS was as valuable, if not more so, than the MPAS answers themselves.

> For some families it’s a good way of opening conversations and also a good way of asking mums to think about their relationship with the baby. And it gives us a way of, you know, mum might after doing the questionnaire have some questions and it gives us a way of exploring that more (HV8)

#### Identifying concerns

Several health visitors reported that MPAS questions were helpful to ‘probe’ understanding of the parent-infant relationship, to identify concerns and to confirm health visitors’ own observations. Some reported that MPAS led to identification of unexpected issues or concerns, such as low mood:

> There was one mum with low mood, but she required listening visits, and I don’t know this little bit of digging [MPAS] brought it to the forefront (HV10)

#### Recording concerns

Some health visitors suggested that MPAS provided a structured way to record parent-infant relationship information. A minority suggested that MPAS was helpful for case management and recording conversations, particularly with parents they had concerns about. One suggested it was useful as a way to evidence clinical practice and demonstrate the worth of health visiting.

> This sort of gives you the opportunity to put it all on paper if you know what I mean and then it’s documented. Whereas we’d just say that we’ve assessed it and that is it. And it was satisfactory or it was good or it wasn’t good, you know. But this sort of gives you that tool where all the questions are there and we’ve done the MPAS and there are no concerns (HV11)

### 4) Challenges of using MPAS in clinical practice

#### Timing and workload

There was a feeling that MPAS was ‘yet another tool’, with other assessments in place that would already pick up concerns, e.g. maternal mood. Health visitors noted that the 3-4 month contact was already information-heavy for parents, and thought that completing the MPAS at this point was burdensome for both themselves and the respondents:

> If you go in and someone’s tired and they’ve got a young baby they don’t want to be doing a form do they? (HV6)

#### Appropriateness of questions

Most of the health visitors commented that some MPAS questions were irrelevant or inappropriate, either due to the timing of asking, or differences in cultural norms. For example, item 9 (“When I have to leave the baby” with response options for feelings of sadness or relief) was not relevant to some mothers at 3-4 months who had not had time away from their infant. There was no ‘not applicable’ option or space to provide context to responses:

> Specially a woman who has a colic baby that cries, and cries, and cries… It can be normal for a woman when that baby is just a knot of scream to have a bit of ‘me time’ for a couple of hours and to be quite honest on the answer and say: well actually no I enjoy not being with him for that two hours but when I go back to them and it was lovely. I would find that perfectly normal. (HV3)

Concerns were raised that questions seemed intrusive, especially if friends or relatives are in the room when health visitors ask the questions. Some suggested that mothers could feel their parenting was being assessed or feel afraid their baby would be taken away, so did not answer honestly:

> I think they maybe think that you…you’re like questioning them and they’re gonna get a mark out of a 10 for being good parents. (HV2)

#### Understanding of questions

Many health visitors found that MPAS uses complex or ambiguous terminology, often needing further explanation. Some questions were perceived as emotive and mothers appeared shocked or uncomfortable. For example, in question 15 (about things mothers have had to give up because of the baby), inclusion of the word ‘resent’ in the responses was criticised. Other questions were thought to be open to interpretation in different ways. For example, in question 17 stating that taking care of the baby is a “heavy burden of responsibility” could be considered normal or interpreted in a negative sense. One health visitor related an incident when the meaning of question 14 (about thinking of the baby as ‘my own’) was misinterpreted by a mother, causing alarm that the baby could have been switched.

Response options were also considered confusing, with the difference between options difficult for mothers to understand. Several health visitors mentioned that they adapted the wording to improve understanding:

> Sometimes without thinking you have to change your language. So you might say ‘very frequently, occasionally or almost never’… So then you maybe refresh it to ‘do you feel really proud of your baby, not very proud of your baby, or you’re not proud at all of your baby you don’t ever feel proud’, you know. So you obviously use terminology that people understand a bit better. (HV5)

#### Length and repetition

Completion time varied considerably, depending on the amount of additional explanation, discussion and follow-up required. Most health visitors thought that MPAS was too long and that questions were repetitive:

> It’s 19 questions, I mean if they say ‘no, no, no’ for example or they’ve never had concerns then it’s fine, but if you start elaborating and saying what do you mean, how do you feel, then obviously it starts taking more time, because then you can’t just ask the question and not explore it further. (HV1)

#### Scoring and referrals

Scoring the MPAS was considered complex and time consuming. Health visitors did not report using the MPAS scores to refer mothers to further services either because few had low scores in the range for concern, or due to a perceived lack of appropriate services in the area. Health visitors also reported concerns about the validity of scores, due to skipping questions or concerns that mothers were answering as they felt they should rather than as they really felt.

#### Women with low levels of English competency

For women with low levels of English language competency, MPAS assessments were conducted by a bilingual health visitor using a transliterated version provided by the research team (Urdu) or with an interpreter (nine other languages). Health visitors reported that the complex concepts and terminology were not easily translatable into a readable format in other languages.

> Urdu or a language like that is quite black and white… “resent”, for example, there’s quite a few ways to describe that word. Whereas in Urdu it’s “do you like your baby, or you hate your baby?”. It’s, that’s kind of it, there’s no middle, happy, balance type thing… so it’s quite hard to translate. (HV1)

Working with interpreters posed challenges, including loss of privacy, which may have affected responses to sensitive questions, and additional time requirements. Health visitors also had concerns that interpreters were not familiar with the concepts and felt unsure whether parents had been given an equivalent interpretation.

> There’s times when they’re speaking a bit longer and I’m just thinking “can you just tell me, you know, what you’re saying to her?” You know, what is this discussion about? Because… they’ve got no training on what these questions mean, interpreters. (HV3)

### 5) Suggestions to improve assessment and support of the parent-infant relationship

Health visitors generally felt it was important to include an assessment of the parent-infant relationship in their clinical practice. Some suggested that they would like to continue using a tool to guide this assessment - to open discussion, provide questions and a score:

> This is the first bonding and attachment kind of tool that we’ve had, or I’ve had any experience with. And I do think that we do need something like that as health visitors. Particularly I think because all professionals are becoming more aware of bonding and attachment… So it is something that we do need (HV7)

When asked about continuing using the MPAS specifically, there was a mixed response. Some were enthusiastic, with just one health visitor suggesting they would not change anything about MPAS, apart from its introduction to mothers. Others were concerned due to the reasons outlined above, and most felt that MPAS could be improved.

Several health visitors expressed a preference for a shorter questionnaire, with repetitive items removed and the option to skip non-applicable questions. They also suggested simpler wording and clearer response categories, with more positively framed questions. Some suggested simplifying the scoring system using a Likert scale or a pictorial approach. Several suggested an approach that used more open questions and discussion:

> It needs to be simplified - it doesn’t need things like, very incompetent, moderately incompetent, moderately competent, competent… I get tongue tied before I’ve finished it and I’m almost exhausted with it at the end of this because there’s all these sections following on from the questions. (HV5)

A suggestion was also made to use MPAS only with a subset of women with a concern, identified during clinical practice or through use of a brief screening question:

> I think using something universally is always challenging and difficult. And people’s understanding of things will vary. But for some people this could, say if you noticed something or mum disclosed something, you could use it in terms of ‘actually we’ve got this questionnaire which might help us hone in on where the problems are’. (HV8)

## Discussion

Health visitors play a crucial role in identifying issues in the parent-infant relationship and intervening early. However, there are currently no recommended tools to assess this relationship and guide provision of appropriate support. We explored the use of one potential measure, the MPAS, in universal health visiting in a deprived and ethnically diverse area of the Northern city of Bradford, UK. The qualitative component of the evaluation explored health visitors’ views on the parent-infant relationship and their experience of using MPAS. The findings highlighted that the universal home visiting service provides a unique opportunity to identify parent-infant relationship concerns, and health visitors saw this as an important part of their role. Some health visitors believed observations to be the best method to assess the parent-infant relationship, with questionnaires being difficult to administer effectively, especially if they had not yet established a relationship with a mother. However, some did value the MPAS as a prompt to open conversations, probe concerns, and improve recording. Whilst some thought that the MPAS confirmed their observations, others reported that it helped to identify issues they had not expected to find, although they were not always aware of appropriate services to refer to. Challenges in using the MPAS in clinical practice were reported, including the length of time taken to administer it, the complexity of the language and the intrusiveness of some questions. These were exacerbated when translation was used. Health visitors’ suggestions to improve this assessment included a shorter tool with more user-friendly language.

There were some variations in health visitor conceptualisation of the parent-infant relationship, with some suggesting that measurement was not required in addition to existing mental health tools. Whilst depression and poor parent-infant relationships are frequently comorbid, they are distinct (Brockington et al., 2006).

Many health visitors expressed a preference for observational tools to assess the parent-infant relationship. Although such observational tools do exist, such as the CARE-Index (Crittenden, 1981; Svanberg et al., 2013) there are disadvantages, including high training costs and time-consuming administration, which would be challenging given the context of reduced budgets and capacity in health visiting (Bryar et al., 2017; Glasper, 2017). Concerns have also been raised about the reliability and validity of unstructured observations (Appleton et al., 2013; Kristensen et al., 2017; Wilson et al., 2010). For routine practice and research, there are potential benefits to a structured approach to minimise bias and improve consistency of reporting. Structured tools may also uncover issues that had not been observed - in this pilot, some health visitors reported that they had identified concerns that they had not expected using the structured MPAS.

The study location of Bradford enabled us to explore specific challenges of administering the MPAS tool with mothers with varying levels of education and English language ability. This provided insight that is relevant for areas with diverse populations. Some limitations were identified. Firstly, we tried to recruit health visitors who had not used the MPAS in the pilot period, as well as those who did. However, all the health visitors who opted to take part had used the tool to some extent (some to a very limited extent), which reduced our understanding of reasons for non-use. Secondly, although health visitors’ reflections on mother’s reactions to the tool provided useful insights, it would be useful to also interview mothers to gain further information on acceptability in the next phase of this work.

### The clinical utility of MPAS in practice: combined discussion of quantitative and qualitative findings

The overall approach used in the pilot, with close, collaborative work between the service and evaluation team, had a number of benefits. In terms of methods, partnership working facilitated a study that was both rigorous and practice-driven with consideration of operational questions. The partnership approach between the evaluation team and clinician and clinical managers also facilitated considerable impact from the study on clinical practice. As a result of working together on the pilot, considerable changes were made to local policy and practice for health visitors in Bradford.

Previous research on tools to measure the infant relationship have provided high quality evidence on the psychometric properties, although studies have often not been conducted in real-world settings, limiting understanding of how tools would work in practice. To our knowledge, this is the first real-world study of clinical utility in practice. Other studies have focussed on utility for health visiting but included limited validation of the tool. This qualitative study and its complementary quantitative study (Dunn et al., submitted) provides evidence on the clinical utility of MPAS that is both methodologically rigorous and grounded in real world clinical practice. Together they revealed concerns about the utility of the tool in practice in this community.

Health visitors interviewed felt that the parent-infant relationship was important, and generally considered assessment of the parent-infant relationship to be an important part of practice. However, quantitative findings showed that only half of mothers (52%) of the 833 mothers who had a 3-4-month health visitor contact during the pilot had been offered the MPAS (Dunn et al., submitted). Whilst this showed that they can integrate an assessment of the parent-infant relationship into their practice, the large number of significant challenges identified in interviews is likely to contribute to the relatively low uptake.

The quantitative analysis indicated that the contextual psychometric properties of the MPAS are not robust enough to recommend ongoing use in the Bradford context. The lack of a stable factor suggests that there was no underlying construct that the women in this study were able to relate to the questions. The qualitative findings elucidate the issues specific to the use of the MPAS tool in the real world, including the difficult marking scheme, complexity of language and difficulties understanding and interpreting questions, especially where women had language needs. The multiple and considerable challenges to using the tool led to inconsistency in the way the MPAS was used and limited utility for clinical practice. Some health visitors reported avoiding using the tool with mothers when they had concerns about their understanding, which could exacerbate health inequalities. These help to explain why the MPAS tool did not work well in real-life clinical practice in a diverse, multi-ethnic urban area. Health visitors also highlighted contextual challenges, especially time constraints during appointments, suggesting that any tool introduced to practice needs to be short and user friendly.

Our pilot evaluation has number of implications for policy, practice and research. In Bradford, deprivation, high levels of health needs, language requirements and cultural differences meant that the tool was tested in a challenging context. In this sense, the location is ideal for a pilot as it is important that any tool to measure parent-infant relationship has clinical utility in diverse populations with high levels of deprivation. Key issues highlighted are of relevance to other urban, multi-ethnic and deprived areas.

Assessment and support for the parent-infant relationship is widely recognised as important. Recent UK guidance has identified the need to “develop reliable and valid screening assessment tools for attachment and sensitivity that can be made available and used in routine … settings” (NICE, 2015). The use of a structured tool enhances the reliability and validity of identification of parent-infant relationship concerns, and quantification of needs. The MPAS was developed on a strong theoretical foundation and has been validated in a range of settings. However, this evaluation provides evidence that the use of MPAS without adaptation would not be valid, acceptable, or feasible in clinical practice in an area with high levels of deprivation and ethnic diversity such as Bradford.

## Conclusion and implications for practice and/or further research

Our linked quantitative and qualitative studies aimed to assess the clinical utility of the MPAS when piloted in health visiting services in Bradford. This study collated qualitative insights from health visitors to understand their views on the parent-infant relationship, their experience of using the MPAS, and develop recommendations for future use in practice.

Overall, our collaborative approach with service providers demonstrated the importance of partnership, both in terms of methodological rigour in a real-world setting, and in terms of impact. Health visitors welcomed the opportunity to discuss the parent infant relationship and there were benefits to using a structured tool. However, there were considerable challenges that hinder implementation of the MPAS in a valid and reliable way – both in terms of the complexity and length of the tool itself and the context that health visitors are working in.

Based on the findings from this paper, and Dunn et al (submitted), although there were benefits to piloting the MPAS, there were also challenges. There remains a gap for a robust, valid measure to assess parent-infant relationships in routine practice, at least in Bradford. Based on these findings, we have coproduced a tool with health visitors, service staff and with input from parents, using the learning from this pilot, and are testing it in routine care.

## Supporting information

Interview guide

## Data Availability

Data is stored securely by Born in Bradford at the Bradford Institute for Health Research (BIHR). Interview participants were informed that data would be stored at BIHR and did not consent to data sharing with other organisations. Therefore, on ethnical grounds the qualitative data are not available to other organisations. The interview guide is provided in supplementary information. For further information about access please contact Born in Bradford (https://borninbradford.nhs.uk/contact-us/).

## Ethics

The Health Research Authority has confirmed that our service evaluation study does not require review by an NHS Research Ethics Committee (HRA decision 60/88/81). However, we have adhered to all ethical principles in the conduct of this evaluation study and written informed consent was obtained from all participants prior to qualitative interviews being undertaken.

## Acknowledgements

This study has received funding through a peer review process from the National Lottery Community Fund as part of the A Better Start programme. The funders have not had any involvement in the design or writing of the paper. Authors PKB, ZH, AD, JD and TB were also supported by the NIHR CLAHRC Yorkshire and Humber (www.clahrc-yh.nihr.ac.uk). JD and TB were supported by the NIHR ARC Yorkshire and Humber (https://www.arc-yh.nihr.ac.uk/). The views and opinions expressed are those of the author(s), and not necessarily those of the National Lottery Community Fund, NHS, the NIHR or the Department of Health and Social Care.

