## Supplementary material for "Implementing the Maternal Postnatal Attachment Scale (MPAS) in universal services: Qualitative interviews with health visitors": Interview guide

### INTERVIEW SCHEDULE FOR INTERVIEWING HEALTH VISITORS TO UNDERSTAND THEIR EXPERIENCE OF USING THE MPAS TOOL

**Version 1.1 11/12/17**

#### Introduction

- *If it's okay with you, I will turn on the audio recorder before we start the interview.*
- *Firstly, thank you for agreeing to be interviewed*
- *My name is xxx and I work as a researcher with Born in Bradford. We are conducting interviews to talk to health visitors in the Better Start area about their experience of using the MPAS tool with parents.*
- *Just to clarify, we do not have any working relationships with the researchers who developed the MPAS tool.*
- *The interview can last as long as you like, but we don't expect it to last more than 40 minutes.*
- *We have a number of questions to ask you. We are interested in your opinions, so there are no right or wrong answers – we want to learn about your experiences and views. Please feel free to mention anything that you think is important and has not been covered in our questions.*
- *The interview is completely voluntary and you can choose to stop it at any time, or skip questions that you would rather not answer.*
- *All your answers are confidential and will only be used for the purpose of this study. When we report the findings we may use quotes, but we will always use pseudonyms instead of real names.*
- *Do you have any final questions before we start the interview?*

**First I'd like to ask a question about you and your role as a health visitor**

1. What do you personally think are the most important issues you address in your role as a health visitor?
   *Physical health, emotional and mental health, parenting, child development, safeguarding?*

**Before we start talking about MPAS, I'd like to ask you about your work and views on the parent-infant relationship**

2. What do you understand by the term parent-infant relationship?
   *What do you consider a healthy parent-infant relationship to be?*
   *What do you consider an unhealthy parent-infant relationship to be?*
   *Are there cultural and individual differences?*

3. What do you think parents understand by the term parent-infant relationship?
   *Do you use the term 'parent-infant relationship' with parents? What terms do you use? What terms do parents use?*

4. Have you received any training about what a healthy parent-infant relationship is?
   *What was this training? Who was it delivered by? When was it? Was it useful?*

5. Do you think health visitors can influence parent-infant relationship? If so, how?
   *What have you done when you are concerned about a woman's relationship with their infant?*

**Now I'd like to ask you about how health visitors assess the parent-infant relationship**

6. In your experience, what do you think are the best ways of measuring the parent-infant relationship?
   *Which measures have you used in the past? Why?*
   *Observation? Parent self-report? Professional assessments?*

**Now I'm going to ask you some questions about using MPAS, and the things that helped or hindered the use of MPAS**

7. Have you ever used MPAS to assess the parent-infant relationship?
   *If used - when/how often, why did you use it?*
   *Did you use MPAS with everyone? If not, what was the decision-making process behind choosing to use MPAS?*
   *If never used - why not? Do you ask any other questions in relation to the parent-infant relationship?*

8. [If health visitors have used MPAS] Did you use the screening question ("Do you have any worries about bonding with your baby?" – health visitors are using this question before the MPAS in the pilot)?
   *If used - when/how often, why did you use it?*
   *Did you use the screening question with everyone? If not, what was the decision-making process behind choosing to use the screening question?*
   *If never used - why not?*

9. Thinking first from your point of view as a health visitor, how did you find using the MPAS with women? What was easy about using MPAS? What was difficult?
   *How did you find fitting it in with your other work?*
   *How did you find explaining it and giving it to women?*
   *Did you have training that helped you use MPAS?*

10. Now thinking from the point of view of mums. Do you think they found the MPAS easy to answer?
    *Do you think that mums understand the language and concepts in MPAS? Why?*

*Are any sections particularly easy/difficult for mums to understand?*

11. How did the mums respond to completing the MPAS?
    *How did you introduce the MPAS to women?*
    *Did mums respond in the way you expected them to?*

12. When you had completed an MPAS, did you take any action based on the score?
    *When the score was low what actions did you take?*
    *When the score was not low, did you ever take any actions?*

13. Has using MPAS changed what you do if you are concerned about a parent's relationship with their infant?

**Now I'm going to ask some questions about the future and what you think could be improved.**

14. Would you like to use/continue using MPAS as part of your clinical practice?
    *If yes, why?*
    *If not, why not?*

15. What would be the main things you would change about the way health visitors assess and support the parent-infant relationship? Why?
    *Would you rather use a different tool or methods altogether? Which and why?*
    *Are there any changes you would make to the way you support parents with attachment concerns? What and why?*

**Bringing our interview to a close…**

16. Is there anything else you would like to say about MPAS and the parent-infant relationship?

***NEXT STEPS***

- *Thank the participant and offer a further chance for questions*
- *Reassure the participant about confidentiality and remind them of contact details in the participant information sheet*
- *Discuss with participant that the findings of the study will be used to help BSB develop the right changes in the area and if they have requested resulting reports then assure these will be shared.*
